# Reconstructing long-term dengue virus immunity in French Polynesia

**DOI:** 10.1101/2022.03.31.22273157

**Authors:** Takahiro Nemoto, Maite Aubry, Yoann Teissier, Richard Paul, Van-Mai Cao-Lormeau, Henrik Salje, Simon Cauchemez

**Author notes:** Equal senior contribution (Van-Mai Cao-Lormeau, Henrik Salje, Simon Cauchemez).

## Abstract

**Background:** Understanding the underlying risk of infection by dengue virus from surveillance systems is complicated due to the complex nature of the disease. In particular, the probability of becoming severely sick is driven by serotype-specific infection histories as well as age; however, this has rarely been quantified. Island communities that have periodic outbreaks dominated by single serotypes provide an opportunity to disentangle the competing role of serotype, age and changes in surveillance systems in characterising disease risk.

**Methodology:** We develop mathematical models to analyse 35 years of dengue surveillance (1979-2014) and seroprevalence studies from French Polynesia. We estimate the annual force of infection, serotype-specific reporting probabilities and changes in surveillance capabilities using the annual age and serotype-specific distribution of dengue.

**Principal Findings:** Eight dengue epidemics occurred between 1979 and 2014, with reporting probabilities for DENV-1 primary infections increasing from 3% to 5%. The reporting probability for DENV-1 secondary infections was 3.6 times that for primary infections. Reporting probabilities for DENV-2–DENV-4 were 0.1-2.6 and 0.7-2.3 times that for DENV-1, for primary and secondary infections, respectively. Reporting probabilities declined with age after 14 y.o. Between 1979 and 2014, the proportion never infected declined from 70% to 23% while the proportion infected at least twice increased from 4.5% to 45%. By 2014, almost half of the population had acquired heterotypic immunity. The probability of an epidemic increased sharply with the estimated fraction of susceptibles among children.

**Conclusion / Significance:** By analysing 35 years of dengue data in French Polynesia, we characterised key factors affecting the dissemination profile and reporting of dengue cases in an epidemiological context simplified by mono-serotypic circulation. Our analysis provides key estimates that can inform the study of dengue in more complex settings where the co-circulation of multiple serotypes can greatly complicate inference.

**Author summary:** Characterising the true extent of dengue circulation and the level of population immunity is essential to assess the burden of disease, evaluate epidemic risk and organise prevention strategies against future epidemics. However, this is difficult in a context where most people who are infected by dengue virus (DENV) only have mild symptoms which may not be reported to surveillance systems. In this article, we develop a mathematical model to evaluate the fraction of unreported dengue infections from case data. The key idea is to introduce reporting probabilities that depend on the infecting serotype and the infection history of patients. These factors are known to contribute to variations in the severity of symptoms and hence the reporting probabilities, but have rarely been taken into account in model frameworks to study population immunity from the case data. Using the developed model, we study long-term dengue virus immunity in French Polynesia.

## Introduction

Dengue, which has four serotypes (DENV-1, DENV-2, DENV-3, DENV-4), is the most widespread mosquito-borne virus infection of humans. The vectors that transmit the dengue virus (DENV) are *Aedes* spp. mosquitoes, primarily *Aedes aegypti*, whose habitat is distributed in tropical and subtropical regions throughout the world. There are an estimated 3.97 billion people at risk of infection worldwide (Brady et al. 2012). Most infections result in mild disease symptoms or are inapparent and are thus not reported to surveillance systems. However, certain patients suffer from severe symptoms, such as Dengue Hemorrhagic Fever (DHF) and Dengue Shock Syndrome (DSS) that have mortality rates of more than 20% if not properly treated (WHO 2020). Overall, 40,000 people died of dengue in 2017 (Roth et al. 2018). Understanding risk factors for severe dengue is important to improve the clinical management of patients.

Many studies have shown evidence of higher incidence rates of severe dengue in secondary infected individuals (Halstead et al. 1967; Guzman, Alvarez, and Halstead 2013). Antibodies for the infecting serotype are produced upon infection. *In vitro*, these antibodies are observed to be cross reactive to the other serotypes: when antibody titres are not high enough, they enhance the entry of the other DENV serotypes into the host cells (Halstead and O’rourke 1977; Mady et al. 1991; Dejnirattisai et al. 2010). This phenomenon, called Antibody Dependent Enhancement (ADE), contributes to a severe disease outcome following infection. However, it is still not clear quantitatively and even qualitatively how severity is linked to the serotype, age, and the infection history of the patient (Guzman, Alvarez, and Halstead 2013). Part of the challenge comes from the difficulty to reconstruct the infection history of patients in a context of high cross-reactivity between serotypes and in the common situation where multiple serotypes co-circulate.

Here we aim to characterise how the history of dengue infection of a patient (i.e., when they were infected and by which serotype) may affect disease severity, considering over 30 years of surveillance as well as serological studies from French Polynesia, which consists of 119 islands, located in the middle of the Pacific Ocean (ISPF 2017) with 276 000 inhabitants (2017) (Fig 1a). We develop a mathematical model that describes how the circulation of dengue serotypes over multiple years has shaped the age-stratified immunity profile of the population, which in turn affects the reporting of infections in surveillance systems (Rodriguez-Barraquer, Salje, and Cummings 2019). Under the assumption that reporting depends on disease severity, we use this framework to investigate the key drivers of disease severity and quantify their relative contributions. Compared to other settings such as in Southeast Asia, where multiple serotypes circulate each year, dengue epidemics in French Polynesia were mono-serotypic until only recently (2013-2014), making it possible to confidently reconstruct likely serotype-specific histories of infection, facilitating the evaluation of the impact of different infection histories on disease severity. We also investigate whether statistics that are estimated within our modelling framework, such as the fraction of susceptibles, can be used to anticipate the occurrence of an epidemic.

**Fig.1.**
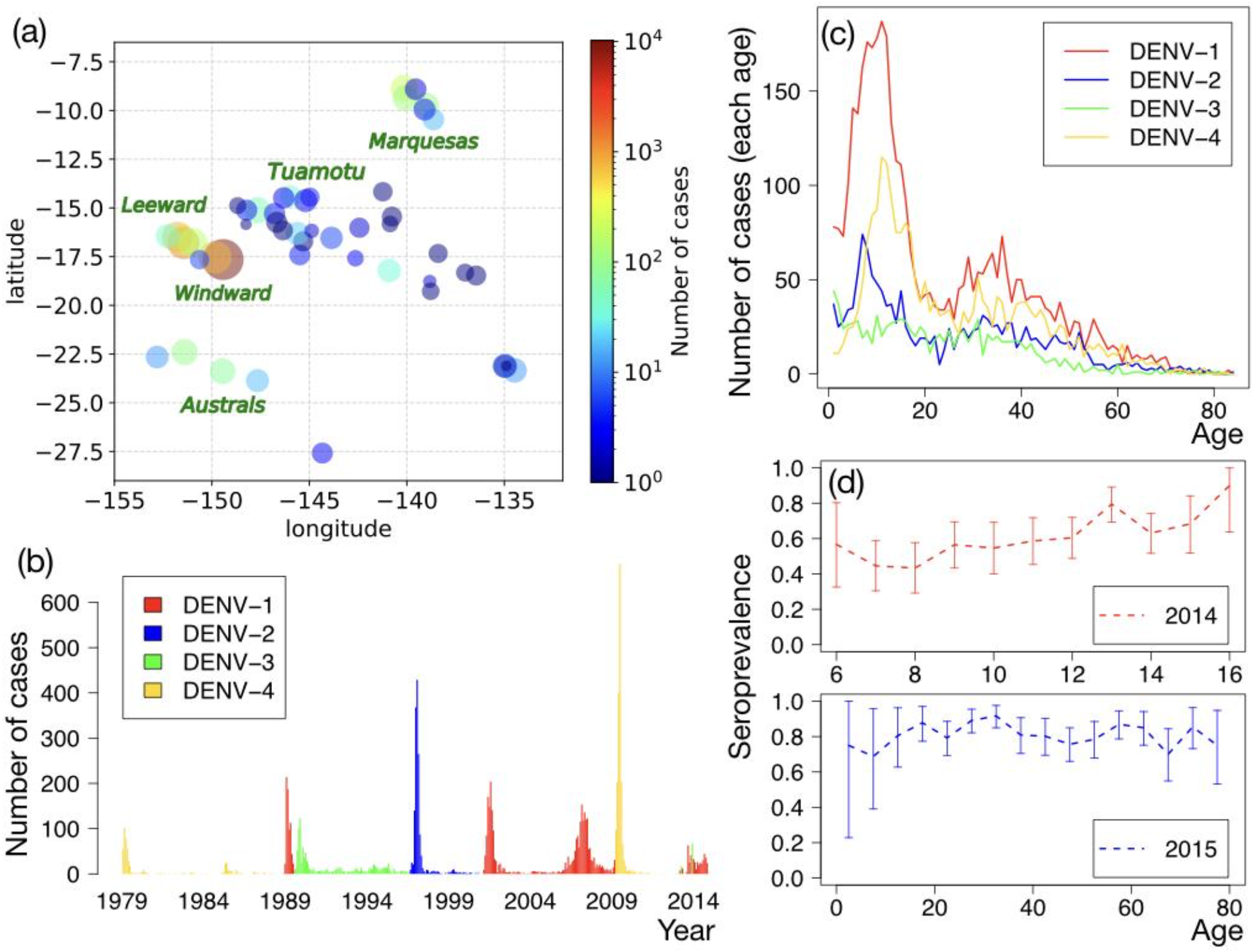
Epidemiology of dengue in French Polynesia. **(a)** Spatial distribution of the number of cases reported between 1979 and 2014, in the different islands of French Polynesia. Each circle represents the size of the population, where the radius is defined as 0.2log_10_P with the population size P. The colours of the circles represent the number of reported cases. **(b)** Monthly number of cases reported for DENV-1 to DENV-4. **(c)** Age distribution of cases, averaged over the period between 1979 and 2014. **(d)** The results of serological surveys (seroprevalence of antibodies against DENV) conducted in 2014 and 2015. Error bars indicate 95%-CI.

## Materials and methods

### Data

#### Surveillance data

The islands constituting French Polynesia are grouped into five administrative subdivisions: Windward, Leeward, Marquesas, Austral, and Tuamotu-Gambier (Fig.1). Since March 1975, Institut Louis Malardé in French Polynesia has received samples of suspected Dengue fever cases over the islands (Teissier et al. 2020). These samples have been analysed with different laboratory techniques, such as Haemagglutination Inhibition Assay (between 1975 and 1988), ELISA-IgM (1986-2003), isolation of DENV on mosquito C6/36 cell culture (1984-2005), and Reverse transcriptase Polymerase Chain Reaction (since 2000). Records include: the results of the tests (positive or negative), the serotype, the age of the patient, and the home community of the patient (from 1978).

In this work, we focus on the most inhabited subdivision, Windward, that includes three populated islands including Tahiti that alone accounts for 70% of the entire population of French Polynesia. We use the data from January 1979 to October 2014 and ignore data for children less than 1 year old to exclude the effect of maternal passive immunity (Halstead et al. 2002). To reconstruct the proportion of the population that were of each age in each year, we use demographic data from the census records of 1971, 1983, 1988, 1996, 2002, 2007 and 2012 (ISPF 2017).

#### Seroprevalence data

Seroprevalence of antibodies against DENV was studied during May-June 2014 and during September-November 2015 in French Polynesia (Aubry et al. 2018). For the study conducted in 2014, 476 school children from primary and highschools in the most populous island (Tahiti) were recruited, while for the study in 2015, 700 people from the general population in the most inhabited subdivision (Windward) were recruited (Fig.1a). The blood samples of these participants were examined using a recombinant-antigen-based indirect ELISA against each serotype in the 2014 study (Aubry et al. 2015b, 2015a) and microsphere immunoassay (MIA) with the same recombinant-antigens used for ELISA in 2015 study (Cao-Lormeau et al. 2016).

### Mathematical model

Our model is composed of the following three parts: a model of dengue circulation in the population, a model of the surveillance system, and a model of serological surveys. We detail these models one by one and combine them in a likelihood function at the end.

#### Modelling dengue circulation and immunity in the population

The Force of Infection (FOI) characterises the transmission intensity of DENV and is defined as the per-capita rate at which susceptible individuals are infected. We denote *λ*_*i*_(*t*) the FOI for serotype *i* (*i* = 1,2,3,4) at time *t*. We assume that the FOI is a step function and define time periods during which the FOI is assumed to be constant, as shown in Supporting information S.2 Table (Note that these time periods correspond to epidemic periods except when a small number of dengue cases is reported. In these cases, the time periods simply correspond to one year.). The j-th time period is labelled *T*_*j*_ (*j* = 1, …, *J*), whose start time and duration are denoted *t*_*j*_ and *Δt*_*j*_, respectively. Since epidemics are mono-serotypic (Teissier et al. 2020), there is only one serotype with a non-null FOI at any given time. Data are also available before 1979 but they are more limited (Aubry and Cao-Lormeau 2019). They include the epidemic period and the circulating serotype. We thus set *λ*_*i*_(*t*) before 1979 to be non-zero only during the epidemic periods with the serotype *i*, where the magnitude of *λ*_*i*_(*t*) (if it is not zero) is constant across epidemics.

Using *λ*_*i*_(*t*), the immunity of the population is derived as follows. We denote by *x*(*t, a*) the fraction of the population at age *a* (*a* = 1,2, …) that has never been infected by dengue before *t*. This population is susceptible to all dengue serotypes. *x*(*t, a* | *λ*) is computed as (Ferguson, Donnelly, and Anderson 1999)

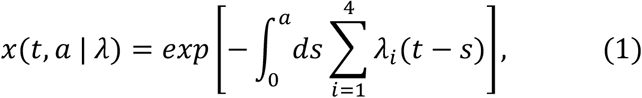

Similarly, we denote by *y*_*i*_(*t, a*) the fraction of population at age *a* who has been infected once by serotype *i* before *t* but is still susceptible to other serotypes (Ferguson, Donnelly, and Anderson 1999):

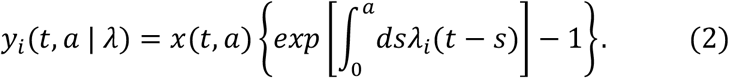

We then denote by *z*(*t, a*) the fraction of population at age *a* who has been infected more than once before time *t*. We assume that this population is not susceptible to any dengue serotype based on (Gibbons et al. 2007) that showed tertiary and quaternary infections are negligible. Note that 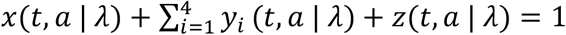. The framework to compute these fractions of population based on FOI is known as the sero catalytic model and has been used in many studies (Schenze, Dietz, and Frösner 1979; Grenfell and Anderson 1985; Ferguson, Donnelly, and Anderson 1999; Kretzschmar, Teunis, and Pebody 2010; Miller et al. 2010; Rodriguez-Barraquer et al. 2011; Bretscher et al. 2013; Muench 2013; Imai et al. 2015; Salje et al. 2016).

Denote 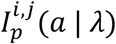 and 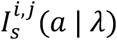 the number of individuals newly infected by serotype *i* (*i* = 1,2,3,4) at age *a* during time period *T*_*j*_ for primary and secondary infections, respectively. These are computed as

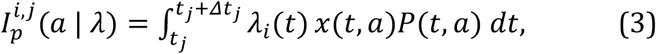

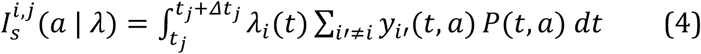

for *i* = 1, …, 4, where *P*(*t, a*) is the number of individuals aged *a* at time *t*. Practically, to evaluate these integrals (and the integrals appearing below), we use the Euler method with step size 4. Assuming that there are no tertiary infections (and higher), the total number of newly infected individuals at age *a* during time period *T*_*j*_ is given as 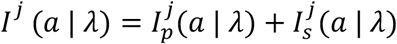, where 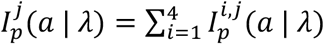 and 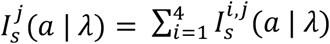.

Finally, we consider the effect of cross protection (Reich et al. 2013) for secondary infections. Upon primary infection, antibody levels rise and then slowly reduce over time (Salje et al. 2018). It was estimated that antibodies cross-protect the patient from other DENV serotypes for an average of 2 years after the primary infection, followed by a period when antibodies may cause ADE (Reich et al. 2013). We assume that cross protection does not alter the immune profile of the population, *x*(*t, a* | *λ*) and *y*_*i*_(*t, a* | *λ*), but only affects the detected number of infected people (*i*.*e*., we assume that those who are protected become asymptomatic, when they are infected, due to cross protection). We denote *δx*_*i*_(*t, a*) the fraction of population aged *a* at time *t* who has been infected by DENV-*i* as primary infections in the past 2 years. Using this quantity, we replace *y*_*i*._(*t, a* | *λ*) in 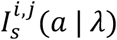 by *y*_*i*._(*t, a* | *λ*) − *δx*_*i*._(*t, a*), which is the true fraction of susceptible for the secondary infections by DENV-*j* (*j* ≠ *i*), excluding those who are protected due to cross protection. (Precisely, some of the individuals who are once in *δx*_*i*_(*t, a*) can be secondarily infected during the 2 years without symptoms. We ignore them by assuming that their contribution is small.) Once taking into account the cross protections, the number of secondary infected individuals of age *a* during time period *T*_*j*_ by DENV-*i* is:

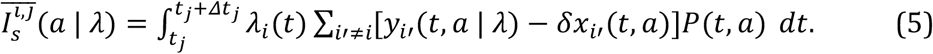

#### Modelling the surveillance system

Not all infected individuals are reported to the surveillance system. Some may be asymptomatic and not seek medical attention. Others may consult a medical doctor and yet not be recommended for a dengue test. Reporting of infected individuals can depend on

1. the age of the individual (infants, adolescents, or adults)
2. the time when the individual is infected
3. the infection history of the individual (primary infection or secondary infection)
4. the infecting serotype (DENV-1, DENV-2, DENV-3, DENV-4)

In order to take into account these different factors, we define the reporting probability *Φ*(*t, a, i, s*) as a function of *t* (the time of infections), *a* (the age of the infected individual), *i* (the infecting serotype (*i* = 1,2,3,4)), and *s* (an indicator of primary (*s* = 1) or secondary infections (*s* = 2)). To reduce the number of free parameters in the model, we assume the following dependency structure for reporting probabilities:

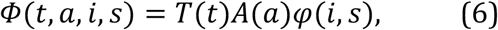

where *φ*(*i, s*) is the relative strength of the reporting probabilities, compared with primary infections of DENV-1 (i.e., *φ*(1,1) = 1). We assume that the age factor *A*(*a*) is constant over age groups 1-4 y.o., 5-9 y.o., 10-14 y.o., 15+, where we set the value for 5-9 y.o. at 1. Finally, the time factor *T*(*t*) is assumed to be constant during the following time periods 1979-1985, 1986-1990, 1991-1995, 1996-2000, 2001-2004, 2005-2008, 2009-2014, and increase between them. *T*(*t*) is the reporting probability for *i* = *j* = 1 and for the age group of 5-10 y.o..

The predicted number of reported cases of age *a* during time interval *T*_*j*_ is modelled using negative binomial distributions with mean

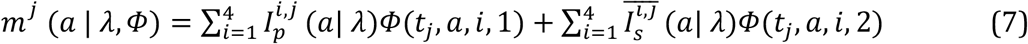

and dispersion parameter

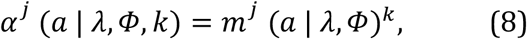

where *k* is a fitting parameter. (Note that, using the average and the dispersion parameter, the variance is expressed as *m* + *m*^2^/*α*. In the large *k* limit, the variance converges to *m* and the distribution becomes the Poisson distribution.)

Our model of the surveillance system is based on (Rodriguez-Barraquer, Salje, and Cummings 2019). Technically, the differences from the previous work are that (i) the reporting probabilities in our model fully characterise how reporting probabilities vary with serotype, age, and the infection history of the patients, and (ii) the previous work used the Poisson distribution for the prediction, which led to narrow confidence intervals that may not reflect underlying statistical uncertainty, while we use the negative binomial distribution, from which broader confidence intervals can be produced.

#### Modelling serological survey

For the *m*-th serological survey, we denote by *t*_*m*_, *n*_*m*_(*a*), and *C*_*sero,m*_(*a*) the time when the survey was performed, the number of participants of age *a*, and the number of participants of age *a* with antibodies (*t*_1_ = 2014.42 and *t*_2_ = 2015.75). From the reconstructed immune profile of the population, the seroprevalence of antibodies against any dengue serotypes *S*(*t*_*m*_, *a* | *λ*) is calculated as

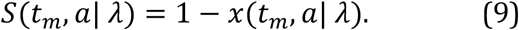

We assume that the probability that the participants have the antibodies follows a Binomial distribution with the number of trial *n*_*m*_(*a*) and a success probability *S*(*t*_*m*_, *a*| *λ*).

#### Likelihood function and Bayesian inference

We denote by 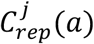 the number of reported cases for age *a* and for time interval *T*_*j*_. The likelihood function *L*(*C*_*rep*_, *C*_*sero*_|*λ, Φ, k*) is the product of the contributions from case reporting and from serological surveys:

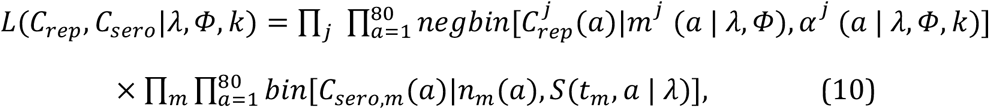

where *negbin*[*C*|*x, y*] is the negative binomial distribution function of *C* with the mean *x* and the dispersion parameter *y, bin*[*C*|*x, y*] is the binomial distribution function of *C* with the number of trial *x* and a success probability *y*. We use a Bayesian Markov Chain Monte Carlo (MCMC) framework and fit the model to the data. We determine the parameters *λ, Φ, k* using a uniform prior. We perform 450 000 iterations with 30 000 iterations as burn-in. We define a 95% credible interval using 2.5% and 97.5% percentiles of the posterior distribution. See Supporting information for more details of this inference and visualisations of the results.

#### Evaluation of the inferential framework using synthetic data

In order to evaluate the accuracy of our Bayesian inference to infer the model parameters, we first generate synthetic data by using the model described above. The model parameters generating these data are chosen as the median of the inferred parameters from the real data. We then infer the model parameters for the synthetic data using our inferential framework. By comparing the true model parameters with the inferred ones, we evaluate the accuracy of our Bayesian inference.

## Results

### Epidemiology of dengue in French Polynesia

The spatial distribution of the reported cases is reported in Fig. 1 (a). It shows that the reported cases are concentrated in Windward islands. Eight epidemics occurred during the 35 years between 1979 and 2014 (Fig. 1 (b)). Note that, in French Polynesia, mono-serotype epidemics had been recorded since 1944 until the recent heterotypic outbreak 2013/2014. Until the emergence of Zika in 2013, the only arboviruses to be identified as circulating were the DENV serotypes (Cao-Lormeau et al. 2014). Children representing the majority of dengue cases (Fig. 1 (c)). The seroprevalence of antibodies against DENV is around 0.8 for the general population while it is around 0.6 for children, indicating children are more susceptible to DENV (Fig.1(d)).

### Reporting probabilities

Under the reasonable assumption that the surveillance system improves over time, we estimate that, in French Polynesia, the probability of reporting a primary infection by DENV-1 increased from 3.13% in 1979 (95%-CI 2.09%-4.39%) to 5.10% in 2014 (95%-CI 3.86%-7.15%)(Fig 2a). The reporting probability for secondary infections by DENV-1 was 3.58 (95%-CI 2.20-5.38) times larger than that for primary infections by the same serotype (Fig. 2(b)), reflecting a general result that secondary infections are more likely to be reported due to increased symptom severity (Guzman, Alvarez, and Halstead 2013). Fig. 2(c) shows how the reporting probability changes with the serotype and whether it is a primary and secondary infection, considering DENV-1 as the reference serotype. For primary infections, reporting probabilities for DENV-2, DENV-3 and DENV-4 are 0.82 (95%-CI 0.51-1.38), 2.61 (95%-CI 1.35-5.71) and 0.11 (95%-CI 0.05-0.23) times larger than that for DENV-1. For secondary infections, the reporting probabilities are 0.69 (95%-CI 0.48-1.08), 2.26 (95%-CI 1.33-4.31) and 0.94 (95%-CI 0.64-1.51) times larger for DENV-2, DENV-3 and DENV-4 respectively than the one for a secondary DENV-1 infection. Note that the reporting probabilities for DENV-4 are small for primary infections. This is in line with observations in Thailand (Salje et al. 2017, 2018) and Nicaragua (Montoya et al. 2013), where DENV-4 was shown to be circulating even though there were not many reported cases of that serotype. We also found that cases older than 14 years old are 0.45 (95%-CI 0.37-0.58) less likely to be reported than those aged 5-9 year old (Fig. 2(d)). Reporting probabilities for children aged 1-4 y.o. and 10-14 y.o. are of the same order as those for children aged 5-9 y.o.

**Fig.2.**
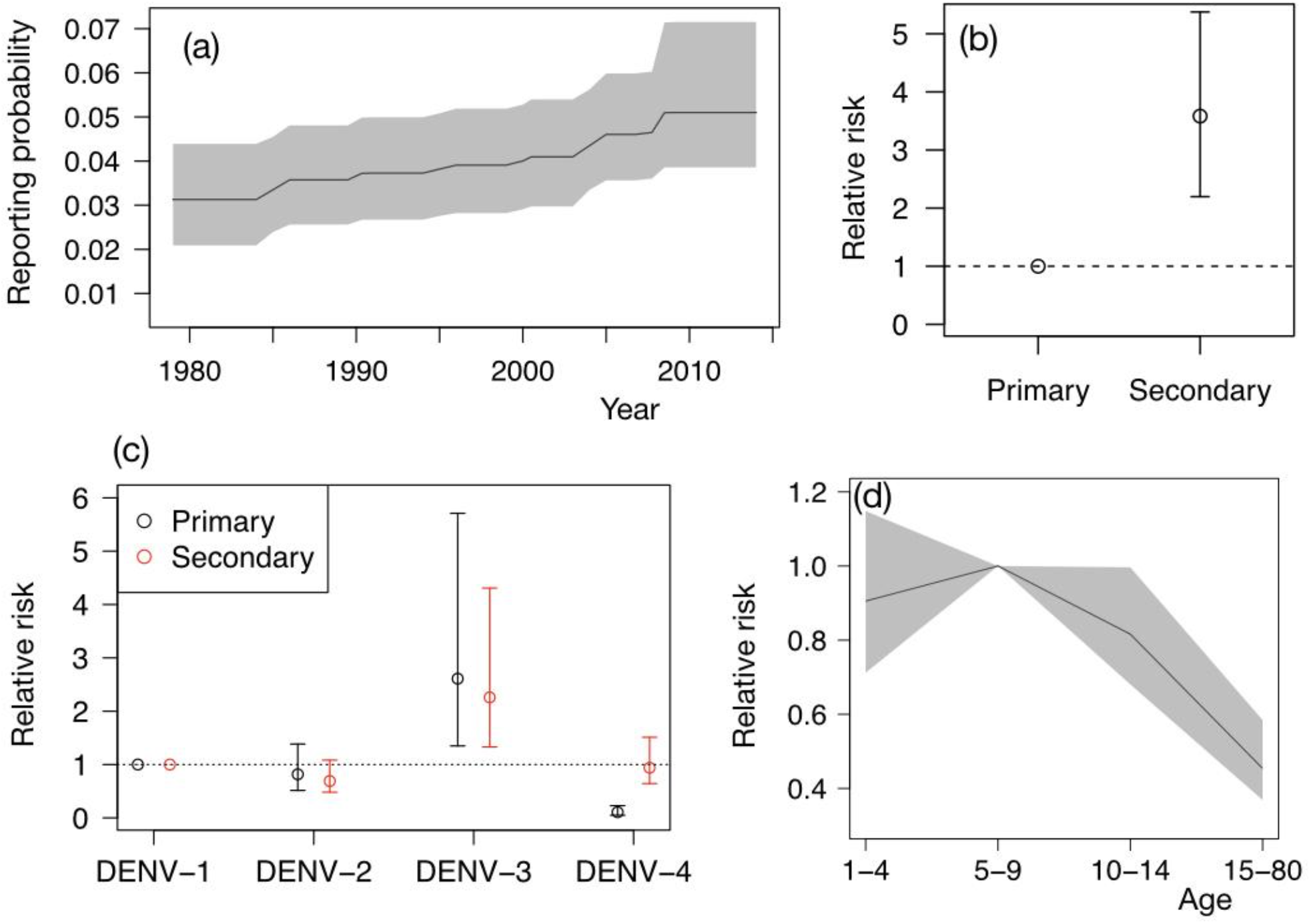
Estimated reporting probabilities. **(a)** Reporting probability of primary infections by DENV-1 as a function of time. The grey shaded area shows 95%-CI. **(b)** Relative strength of the reporting probabilities of secondary infections (DENV-1) compared with primary infections (DENV-1). **(c)** Comparison of the reporting probabilities for different serotypes, for primary (black circle) and secondary (red circle) infections. The reference group is primary Serotype 1 infection for primary infections and secondary Serotype 1 infection for secondary infections. **(d)** Variations of the reporting probability with the age group, considering individuals aged 5-9 year old as the reference group.

### FOI and immunity in the population

Next, we compare the observed and expected number of reported cases per year in Fig.3(a), while estimated FOI are presented in Fig.3(b). The FOI and the number of reported cases show qualitatively similar peaks, but some details are different. For example, the largest epidemics in terms of reported cases occurred in 2009 and was due to DENV-4 (2853 / year), followed by the second largest in 1996 due to DENV-2 (1983 / year). In terms of FOI, the extent of these two epidemics is more or less the same, with an FOI at 0.65 (95%-CI 0.44-0.85) for the 2009 DENV-4 epidemic and 0.59 (95%-CI 0.43-0.76) for the 1996 DENV-2 epidemic. This could be because (i) the quality of surveillance systems improved over time as seen from Fig.2(a), and also because (ii) the secondary-infection reporting probabilities for DENV-2 (relative risk: 0.69) are smaller than those for DENV-4 (0.94) and DENV-1 (1). The same argument can also explain the relatively small FOI observed for DENV-1 in the epidemics occurring after 2000.

**Fig.3.**
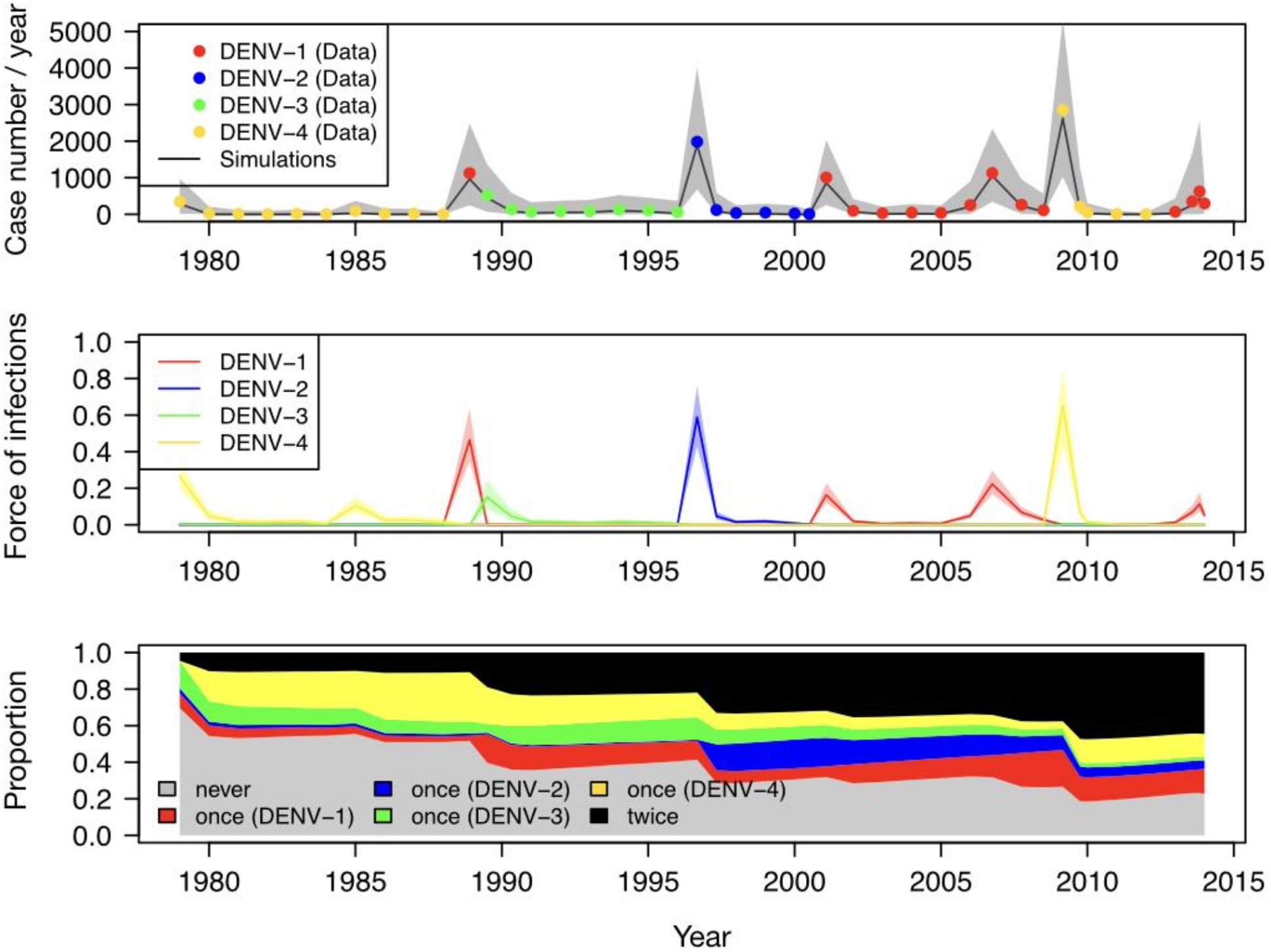
Estimated FOI and immunity. **(a)** The observed (dots) and expected (line) number of cases reported annually. Shaded area represents 95%-CI. **(b)** Fitted FOI for the four serotypes. **(c)** Average immunity profile of the population. The grey area shows the fraction of the population that were never infected, averaged over age groups. Red, blue, green, and yellow areas represent the fraction of the population who have been infected once by a serotype i (before the time we consider), where i = 1,2,3,4 correspond to red, blue, green, yellow, respectively. Black area represents the fraction of the population who have been infected more than once (before the time we consider).

Using the estimated parameters, we reconstruct the immunity profile of the population during the surveillance period and plot it in Fig.3(c). The graph shows that the immunity profile changed over time. During the course of the surveillance period (1979-2014), the proportion that were never infected declined from 0.70 to 0.23 while the proportion that were at least twice infected increased from 0.045 to 0.45 at the end of the period (2014). The general population has experienced several dengue epidemics during the 35 years of surveillance. As a result, almost half of the population has acquired heterotypic immunity.

### Age distribution and immunity

In Fig.4(a), we show the age distribution of the number of reported cases for all epidemics during the surveillance period. Overall, the data points fall inside the 95%-CI, showing that our model captures well the age structure of the immunity of the population. The shapes of certain distributions in Fig.4(a) can be qualitatively understood based on the severity of secondary infections as follows. The age distributions of the epidemics during 1988-1989, 1996-1997, 2001-2002, and 2009 have a plateau (or a peak) and to the left side of this plateau, a relatively small number of reported cases is observed. The plateaus start around 10, 6, 5, and 8 years of age for the epidemics during 1988-1989, 1996-1997, 2001-2002 and 2009, respectively. The children whose ages are less than these plateau ages did not experience the previous epidemic. This means that they had less risk of having secondary infections, so that it is consistent with the observation that the age group to the left side of the plateau had less reported numbers. In Fig.4(b), we compare the results of the serological survey (the seroprevalence of antibodies against DENV in 2014 and 2015) (Aubry et al. 2018) with the model predictions using the fitted parameters. For the 2014 survey, most of the model predictions fall within 95%-CI, while for the 2015 survey, small systematic deviations in some age groups are observed.

**Fig.4.**
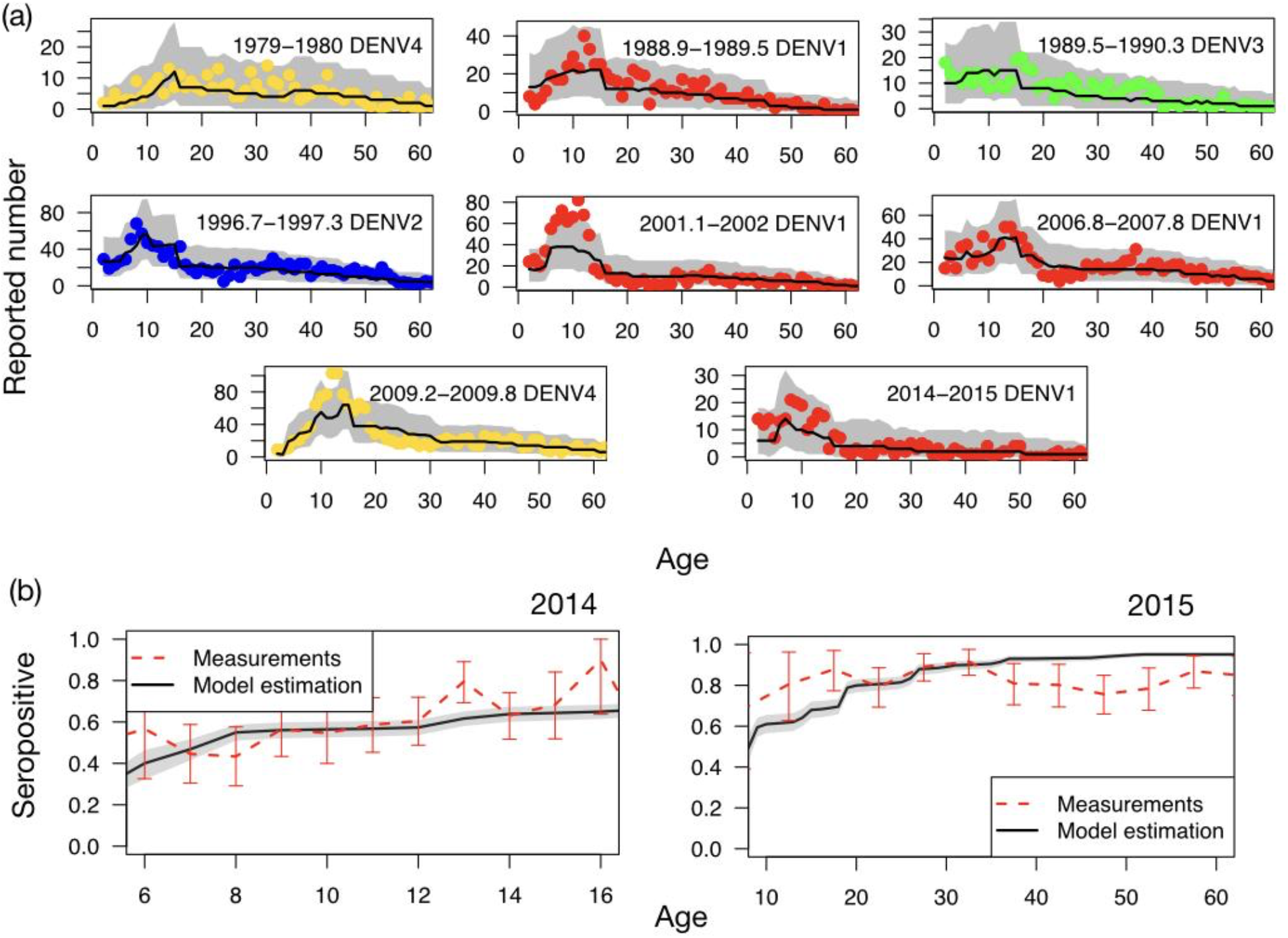
Observed and expected age distribution. **(a)** Age distributions of the reported case numbers during the epidemic periods (red, blue, green, yellow circles correspond to the serotype 1, 2, 3, 4, respectively). **(b)** The seroprevalence of antibodies against DENV obtained from the serological survey (red dashed lines with error bars). In both (a) and (b), black solid lines give model predictions, with 95%-CI represented by the grey shaded areas (95%-CI)

### Relation between the proportion of susceptibles and FOI

In Fig.5(a), we plot the FOI as a function of the fraction of susceptibles among children (left panel) and in the general population (right panel). Child susceptibility has stronger correlation with the FOI (Pearson Correlation Coefficient: 0.65; 95%-CI 0.44-0.80) than adult susceptibility (0.37; 95%-CI 0.08-0.60).

We next estimate the probability of occurrence of an epidemic as a function of the susceptible fraction by using logistic regression. Probabilities are plotted in Fig.5(b) for children (left panel) and for the general population (right panel). The probability sharply increases when the fraction of susceptibles among children exceeds 0.8. This suggests that this indicator might be used as an indicator for the occurrence of epidemics. To study the ability to predict an epidemic based on the fraction of susceptibles, we plot the Receiver Operating Characteristic (ROC) curve in Fig.5(c), where True Positive Rates (TPR) are plotted as a function of False Positive Rates (FPR) for various threshold values for the fraction of susceptibles. We find that the closest point to the perfect classification (FPR=0, TPR=1) is the one using the threshold value 0.82-0.84, leading to FPR=0 and TPR=0.86.

**Fig.5.**
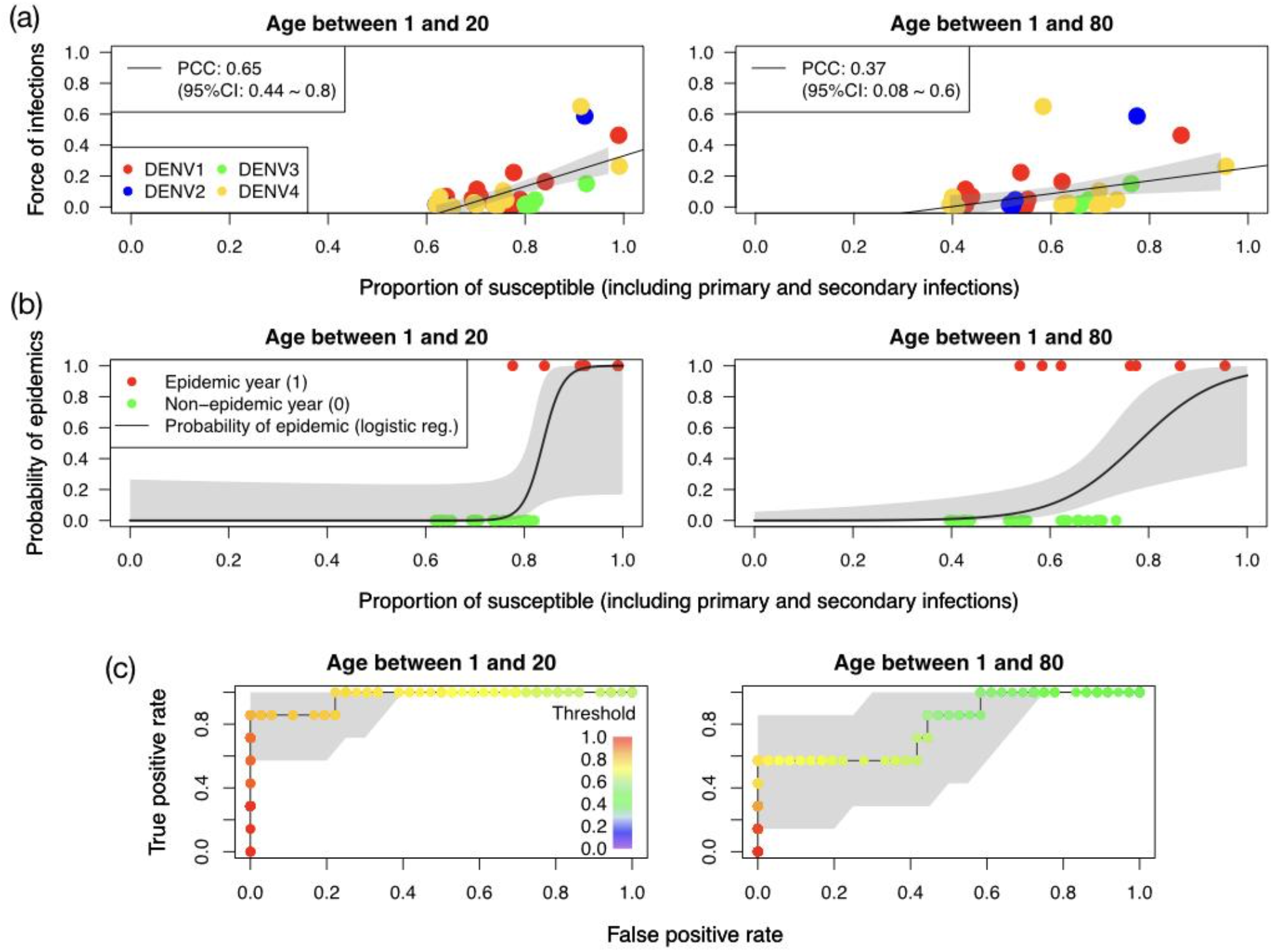
Relation between the proportion of susceptibles and FOI. **(a)** FOI as a function of the fraction of the susceptibles to primary and secondary infections for children (left panel) and general population (right panel). Different colours represent a different circulating serotype. The black solid lines represent the linear regression to the data with 95%-CI as grey shaded areas. Pearson correlation coefficient (PCC) between the FOI and the fraction of the susceptible is also provided in the figure. **(b)** The probability of occurrence of an epidemic (black solid lines) as a function of the fraction of susceptibles. This probability is estimated using application of the logistic regression to the data (See Method Section). Grey shaded areas show 95%-CI. Red and green circles show epidemics and non-epidemic time periods, respectively. **(c)** ROC curve to illustrate the diagnostic ability of predicting an epidemic using the fraction of the population that are susceptible.

### Results for the evaluation of the inferential framework using synthetic data

We evaluate our inferential framework using the method described at the end of the Mathematical model section. In Fig. 6, we show the true parameters and the inferred results for the synthetic data with 95%-confidence intervals. We observe that most of the true parameters are within the credible intervals (95.38%: 62 parameters out of 65), demonstrating the accuracy of our inferential framework.

**Fig.6.**
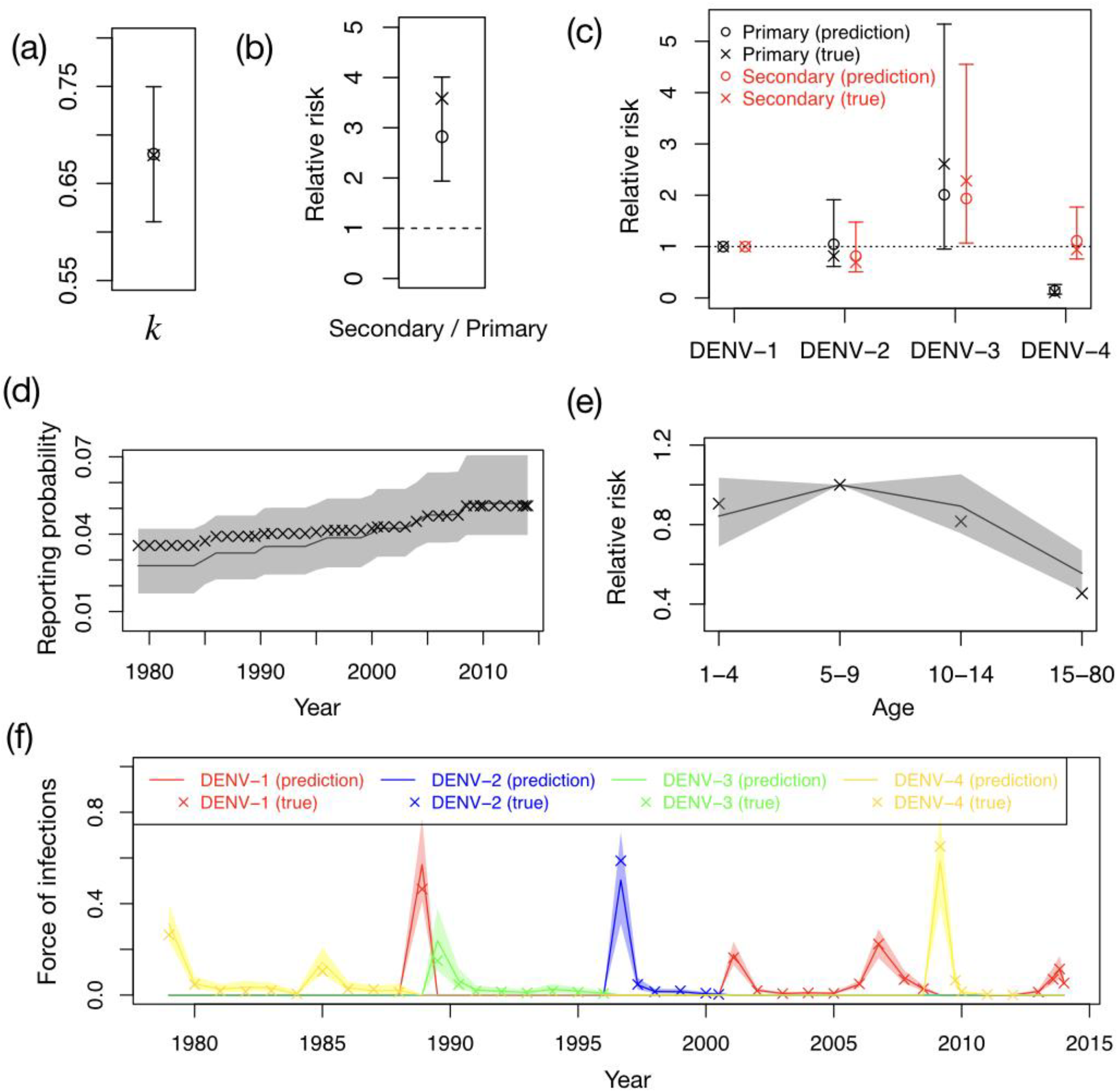
Evaluating the Bayesian inference using synthetic data. Using the model described in the main text, we generate synthetic data for given model parameters. Using this synthetic data, we then infer the model parameters using the Bayesian inference. The original parameters are plotted as crosses (true), while the inferred results are plotted as lines or circles with confidence intervals (predictions). In (a), the parameter k for the negative binomial distribution is shown. In (b), the relative strength of the reporting probabilities of secondary infections (DENV-1) compared with primary infections (DENV-1) φ(1,2)/φ(1,1) is shown. In (c), the reporting probabilities relative to serotype 1, φ(i, 1)/φ(i, 1) for primary infections and φ(i, 2)/φ(i, 2) for secondary infections, are shown. Here i = 1,2,3,4 corresponds to DENV-i. In (d), the reporting probability of primary infections by DENV-1 T(t)is shown as a function of time. In (e), the age-factor of the reporting probability A(a) is shown. In (f), the FOI is shown as a function of time.

## Discussion

In this article, we studied how the reporting probabilities of dengue infections depend on serotype, age, and the infection history of patients. To this goal, we generalised the sero-catalytic model with a reporting structure (Rodriguez-Barraquer, Salje, and Cummings 2019) by introducing reporting probabilities that vary with serotype, age, and the infection history of the patients. We fitted this model to case data in French Polynesia between 1979 and 2014 (Teissier et al. 2020), where only mono-serotype circulations had been observed until the recent heterotypic outbreak 2013/2014, and to the result of the seroprevalence surveys conducted in 2014 and 2015 (Aubry et al. 2018).

The results show that for DENV-1, the reporting probability for secondary infections is about three times higher than for primary infections. We estimated reporting probabilities for different serotypes and showed that DENV-3 infections were the most likely to be reported both for primary and secondary infections. In French Polynesia data, the DENV-3 epidemic and endemic transmission (1990-1996) happened just after the DENV-1 epidemic (1989). This sequence of events, DENV-1 (or DENV-2) followed by DENV-3, showed the highest severity in the Cuban epidemics (Alvarez et al. 2006; Guzman et al. 2012), consistent with our observations. The reporting probability for DENV-2 is similar to the one for DENV-1. This is in contrast to the observation that the secondary DENV-2 infection was 5-7 times more frequently associated with DHF than was secondary DENV-1 or DENV-3 infections in Cuba (Guzman et al. 2012), although the sequence of events was DENV-3 followed by DENV-2 in our case, while it was DENV-1 followed by DENV-2 in the other case (Guzman et al. 2012).

The epidemic of DENV-1 that took place in 2001 was followed by endemic circulation of DENV-1 for 5 years (Teissier et al. 2020). This then led to a new epidemic of DENV-1 in 2006. In the time-series analysis of the case data in different subdivisions (Teissier et al. 2020), it was shown that DENV-1 circulated in the other subdivisions after the first epidemic, and the second epidemic in Windward islands was triggered because of the re-introduction of the virus from the Marquesas. Since our model only considers the Windward islands, we could not detect these inter-subdivision transmissions. Future studies could incorporate the spatial structure of the data and describe these inter-subdivison transmissions. We showed that the shape of the age distribution in Fig.4 was influenced by the year of previous epidemics. The epidemic in 2009 however needs further explanation because the plateau starts at 8 years old, even though, according to this argument, it should start at 3 years old as the previous epidemic by DENV-1 was in 2006. This indicates that those who drove the epidemic in 2006 were not small children less than 5 years old, but slightly older children, who lived through the 2001 epidemic.

In our model, we assumed that only a single serotype circulates at a given time. This assumption was confirmed before the 2013/2014 DENV outbreak (Teissier et al. 2020). To analyse the recent data after this outbreak (and also hyperendemic regions other than French Polynesia), it would be necessary to take into account the concomitant circulation of different serotypes in our model. It would be also interesting to consider the seroprevalence of antibodies against Zika virus, which first emerged in French Polynesia in 2013 (Aubry et al. 2018). As there was no Zika circulation before this period, the data and our model could provide a fruitful test ground to study cross reactivity between Zika and DENV (Dejnirattisai et al. 2016; Priyamvada et al. 2017).

Finally, the currently available licensed dengue vaccine, Dengvaxia, developed by Sanofi Pasteur, is recommended to be used on individuals who have already been infected by one of DENV serotypes (WHO 2020); otherwise, the vaccination can increase the risk of severe symptoms because of ADE (Sridhar et al. 2018). By estimating the seroprevalence, our method could help to identify the region and also the age groups to be vaccinated in the presence of cross reactivities (Rodriguez-Barraquer, Salje, and Cummings 2019) as the WHO’s scientific advisory group of experts committee recommend the use of Dengvaxia only in places with the seroprevalence greater than 70% (WHO 2016). A new tetravalent dengue vaccine developed by Takeda (TAK-003) has been in phase 2 and 3 trials. The phase 2 trial involved the follow up of 1800 participants for 48 months and showed the absence of severe symptoms due to the vaccination of naive individuals (Tricou et al. 2020). There are still some concerns related to the efficacy of this vaccine against DENV 3 that might depend on the infection history of the individuals (Wilder-Smith 2020). An additional 3-year period for long-term efficacy and safety evaluation is being conducted.

By analysing 35 years of dengue data in French Polynesia, we characterised key factors affecting the dissemination profile and reporting of dengue cases in an epidemiological context simplified by mono-serotypic circulation. Our analysis provides key estimates that can inform the study of dengue in more complex settings where the co-circulation of multiple serotypes can greatly complicate inference.

**S1 Appendix. Visualisation of the inferred parameters**.

**S2 Table. Time periods where the force of infection takes a constant value**.

## Data Availability

The data underlying the results presented in the study and the source code will be available from Zenodo.

## Acknowledgments

This project has received funding from the Investissement d’Avenir program, the Laboratoire d’Excellence Integrative Biology of Emerging Infectious Diseases program (grant ANR-10-LABX-62-IBEID), the European Research Council (No. 804744) and the European Commission Seventh Framework Program [FP7/2007-2013] for the DENFREE project under Grant Agreement no.282378.

## Ethics approval

Ethical approval for the data used in this study was obtained from Comité d’éthique de la Polynésie française (Avis Numéro 74 CEPF du 04/05/2018).

## Supporting information

### S1. Visualisation of the inferred parameters

First of all, 95%-CIs used in this article are generated under the Bayesina framework using the negative binomial likelihood probability with the fitting parameter *k* that is inferred as 0.68 (95%-CI 0.61-0.75).

In Fig.2(a), the relative strength of the reporting probabilities of secondary infections (DENV-1) compared with primary infections (DENV-1) is plotted. This is estimated as *φ*(1,2)/*φ*(1,1). In Fig.2(b), the reporting probabilities relative to serotype 1 are plotted for both primary and secondary infections. These are estimated as *φ*(*i*, 1)/*φ*(1,1) for primary infections and *φ*(*i*, 2)/*φ*(1,2) for secondary infections (*i* = 1,2,3,4). In Fig.2(c), the time-dependent factor of the reporting probabilities *T*(*t*) in Eq. (6) is plotted. In Fig.2(d), the age factor of the reporting probabilities *A*(*a*) in Eq. (6) is plotted.

In Fig.3(b), the FOI *λ*_*i*_(*t*) is plotted (*i* = 1,2,3,4 corresponds to DENV-1, DENV-2, DENV-3, DENV-4). In Fig.3(c), the average immunity profile of the population is plotted using the fitted parameters. The fraction of the never infected population averaged over age at the time *t* is estimated as 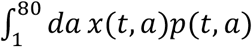, where *p*(*t, a*) is the normalised age distribution of the population size. The averaged fraction of the population who have been infected once by a serotype *i* before the time *t* is estimated as 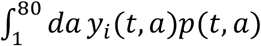. Finally, the averaged fraction of the population who have been infected more than once before the time *t* is estimated as 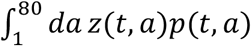.

In Fig.5(a), FOI is plotted as a function of the fraction of the susceptible. Using the fitted model, this fraction (for circulating serotype *i*) is estimated as 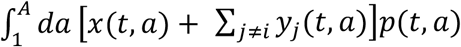 for each time period, where *A* is 20 for the children (the left panel of Fig.5(a)) and 80 for the general population (the right panel of Fig.5(a)). In Fig.5(b), the probability of the occurrence of the epidemic (black solid lines) is plotted as a function of the fraction of the susceptible. This probability is estimated using logistic regression as detailed as follows: We first define the epidemic period as the period during which the reported number exceeds 300. We then plot the value 1 as a function of the fraction of the susceptible for the epidemic periods (red filled circles in Fig.5(b)) and 0 for the non-epidemic periods (green filled circles in Fig.5(b)). Assuming a linear relationship between the fraction of the susceptible and the log-odds of the occurrence of epidemics, we determine this linear coefficient by fitting the corresponding Bernoulli sampling model to the 0-or-1 signal. For the ROC curve in Fig.5(c), we first introduce a threshold value for the fraction of susceptible, above which we expect that the epidemic occurs and below which we do not. We then calculate the FPR (the rate at which an epidemic does not occur even if we expect it to occur) and the TPR (the rate at which an epidemic occurs as expected) for various values of the threshold. In Fig.5(c), we plot filled circles at the position (*x, y*)=(FPR, TPR), where the colours of the filled circles correspond to the threshold value.

**S2.** Time periods where the force of infection takes a constant value.

| Time period | Epidemic | Serotype |
| --- | --- | --- |
| 1 Jan 1979 - 31 Dec 1980 | Yes | 4 |
| 1 Jan 1980 - 31 Dec 1981 | No | 4 |
| 1 Jan 1981 - 31 Dec 1982 | No | 4 |
| 1 Jan 1982 - 31 Dec 1983 | No | 4 |
| 1 Jan 1983 - 31 Dec 1984 | No | 4 |
| 1 Jan 1984 - 31 Dec 1985 | No | 4 |
| 1 Jan 1985 - 31 Dec 1986 | No | 4 |
| 1 Jan 1986 - 31 Dec 1987 | No | 4 |
| 1 Jan 1987 - 31 Dec 1988 | No | 4 |
| 1 Jan 1988 - 24 Nov 1988 | No | 4 |
| 25 Nov 1988 - 2 July 1989 | Yes | 1 |
| 3 July 1989 - 30 April 1990 | Yes | 3 |
| 1 May 1990 - 31 Dec 1991 | No | 3 |
| 1 Jan 1991 - 31 Dec 1992 | No | 3 |
| 1 Jan 1992 - 31 Dec 1993 | No | 3 |
| 1 Jan 1993 - 31 Dec 1994 | No | 3 |
| 1 Jan 1994 - 31 Dec 1995 | No | 3 |
| 1 Jan 1995 - 31 Dec 1996 | No | 3 |
| 1 Jan 1996 - 1 Sep 1996 | No | 3 |
| 2 Sep 1996 - 30 April 1997 | Yes | 2 |
| 1 May 1997 - 31 Dec 1998 | No | 2 |
| 1 Jan 1998 - 31 Dec 1999 | No | 2 |
| 1 Jan 1999 - 31 Dec 2000 | No | 2 |
| 1 Jan 2000 - 1 July 2000 | No | 2 |
| 2 July 2000 - 29 Jan 2001 | No | 2 |
| 30 Jan 2001 - 31 Dec 2002 | Yes | 1 |
| 1 Jan 2002 - 31 Dec 2003 | No | 1 |
| 1 Jan 2003 - 31 Dec 2004 | No | 1 |
| 1 Jan 2004 - 31 Dec 2005 | No | 1 |
| 1 Jan 2005 - 31 Dec 2006 | No | 1 |
| 1 Jan 2006 - 1 Oct 2006 | No | 1 |
| 2 Oct 2006 - 1 Oct 2007 | Yes | 1 |
| 2 Oct 2007 - 1 July 2008 | No | 1 |
| 2 July 2008 - 27 Feb 2009 | No | 1 |
| 28 Feb 2009 - 1 Oct 2009 | Yes | 4 |
| 2 Oct 2009 - 31 Dec 2010 | No | 4 |
| 1 Jan 2010 - 31 Dec 2011 | No | 4 |
| 1 Jan 2011 - 31 Dec 2012 | No | 4 |
| 1 Jan 2012 - 31 Dec 2013 | No | 4 |
| 1 Jan 2013 - 31 July 2013 | No | 1 |
| 1 Aug 2013 - 30 Oct 2013 | No | 1 |
| 1 Nov 2013 - 31 Dec 2014 | No | 1 |
| 1 Jan 2014 - 23 Oct 2014 | Yes | 1 |

## Notes

### Competing Interest Statement

The authors have declared no competing interest.

### Funding Statement

SC and HS have received funding from the Investissement d’Avenir program, the Laboratoire d’Excellence Integrative Biology of Emerging Infectious Diseases program (grant ANR-10-LABX-62-IBEID) https://anr.fr/ProjetIA-10-LABX-0062 and the European Research Council (No. 804744) https://cordis.europa.eu/project/id/804744 RP and VMCL have received funding from the European Commission Seventh Framework Program [FP7/2007-2013] for the DENFREE project under Grant Agreement no.282378 https://ec.europa.eu/research/fp7/index_en.cfm The funders had no role in study design, data collection and analysis, decision to publish, or preparation of the manuscript.

